# PREVALENCE OF PRECONCEPTION CARE PROVISION IN KISUMU COUNTY-KENYA

**DOI:** 10.1101/2022.04.29.22274489

**Authors:** Everlyne N. Morema, Morris Senghor, Collins Ouma

**Author notes:** Corresponding author CO. **E-mail addresses :** ENM, MSS.

## Abstract

**Background:** ‘Preconception care’ (PCC) is the provision of biomedical, behavioral and social health interventions to women and couples before conception occurs. The PCC, is valuable and key in preventing and controlling non-communicable diseases. There is the need for comprehensive preconception care provision in Kisumu County in order to promote healthy reproduction, improve maternal and neonatal health indicators in this and similar settings. Unfortunately, the current rate of provision for this service has not been documented in Kenya and more specifically in Kisumu County.

**Methods:** In a cross-sectional study, data on the provision of various services as per the recommended package for preconception care was collected in health facilities using a checklist. The targeted facilities (n=28) were selected using multistage sampling. The means for all of the services in the package was determined. The significance of the difference in the means was determined by one sample T-test at *P*-value ≤ 0.05

**Results:** Level of implementation of PCC was quite low at 39%. It was observed to be lower in the primary level facilities (KEPH level 2 and 3) at 34% and higher in referral facilities (level 4 and above) at 45%. The service with the highest implementation level was HIV prevention and management (84%) followed by sexually transmitted diseases (80%) and vaccination services (75%). The service with lowest level of implementation was environmental risk exposure reduction at 13% for level 2 and 3 followed by management of mental health disorders.

**Conclusion:** Provision of PCC was relatively low at 39% and provision was fragmented. Provision differed across service levels and care packages.

## BACKGROUND

Preconception care is the provision of biomedical, behavioral and social health interventions to women and couples before conception occurs. It aims at improving their health status, reducing behaviors and individual factors as well as environmental factors that contribute to poor maternal and child health outcomes (WHO, 2013). Preconception cares as an important component of the continuum of maternal health therefore plays a big role in improving maternal and neonatal health indicators (Dean et al., 2013). Preconception care contributes to a healthy pregnancy and a better obstetric outcome (Dean et al., 2014). Preconception care, as part of the national policy framework, is recognized as an important contributor to non-communicable disease prevention and control (Mason et al., 2014; WHO, 2013).

Preconception care has a range of benefits that include;-reduction of maternal and child mortality, prevention of unintended pregnancies, complications during pregnancy and delivery, stillbirths, preterm birth and low birth weight, birth defects, neonatal infections, underweight and stunting, prevent vertical transmission of HIV/STIs, lower the risk of some forms of childhood cancers, lower the risk of type 2 diabetes and cardiovascular disease later in life (Lan et al., 2017; M’hamdi, Van Voorst, Pinxten, Hilhorst, & Steegers, 2017).

Preconception risk factors are well profiled and intertwined. Addressing these factors comprehensively can go a long way in solving much of the problems (Dubei, 2014). Risk factors associated with poor obstetric outcomes like prematurity include; pregnancy in adolescence, birth spacing (short and long intervals) (Shah & Zao, 2009), pre-pregnancy weight status (underweight) (Zhangbin et al., 2013), overweight and obesity (Pirkola et al., 2010), micronutrient deficiencies (iron and folate) (Dean et al., 2014), chronic diseases (DM, hypertension and anemia), poor mental health (depression and intimate partner violence (Alhusen et al., 2014; Alhusen, Lucea, Bullock, & Sharps, 2013; Dean et al., 2013), infectious diseases (mumps, rubella, syphilis and HIV/Aids) (Dean, Imam, Lassi, & Bhutta, 2012), and tobacco use (Bottorff et al., 2014; Caleyachetty et al., 2014; Levine, Cheng, Cluss, Marcus, & Kalarchian, 2013).

Risk factors for unhealthy pregnancy that are amenable to preconception care are present in Kenya and more so Kisumu County. The prevalence of teenage pregnancy and motherhood is remains high in Nyanza region as per the two previous Kenya Demographic Health Surveys (KDHS) (Kenya National Bureau of Statistics et al., 2015; KNBS, 2010). According to these surveys, the median age at first birth is lowest in Nyanza at 18.7 years; birth interval of less than 18 months is common in Nyanza Region at 7.6 per 1000 live births, second only to North Eastern. Kisumu County is at the lead at 8.8 for the same within the Nyanza region. Success in achieving reproductive intention is lowest in Nyanza. The gap between wanted and unwanted fertility is widest in Nyanza at 1.5 in as compared to the national gap which shows women having 0.6 child more than the number they intend to get. The proportion of overweight women of reproductive age has increased significantly from 25-33% since 2008 (Kenya National Bureau of Statistics et al., 2015; KNBS, 2010). This emphasizes on the need for comprehensive preconception care provision in order to promote healthy reproduction, improve maternal and neonatal health indicators in this and similar settings.

The rate of preconception care is low worldwide worse in low-income countries since it is not a wide spread concept as of yet (Lassi, Dean, Mallick, & Bhutta, 2014). The rate of use of these services was found to be between 18.7% and 45% among diabetes patients offered the service in a study done in Canada (Kallas-Koeman, Khandwala, & Donovan, 2012). Another study in England estimated it at 45% (Tripathi, Rankin, Aarvold, Chandler, & Bell, 2010). In United States of America, two studies estimated it at 33% and 47.7% (Bright & Dipietro, 2019; Shadab, Nekuei, & Yadegarfar, 2017). Studies have shown that despite being offered this service very few couples agree to be recruited to the care (Dean, 2014). The studies in Canada, USA and England showed that socio economic status, previous pregnancies and age influenced the uptake of this particular service (Kallas-Koeman et al., 2012; Shadab et al., 2017; Tripathi et al., 2010). This raises the question of what influences its uptake in Kenya.

Preconception care interventions in various settings particularly in low-income countries face the challenge of lack of standardization across the line of service delivery, community outreach and organizational policies (Dean et al., 2014). Preconception care still remains a new concept because it is not included in the Kenyan maternal health model as well as the Kenyan health strategic plan. This has minimized the focus on this service thus little is known about the rate of its provision and use and factors that influence it.

Sustainable development goal three highlights the fact that only half the women in developing countries have received the health care they need. Therefore, one of its targets is to reduce maternal mortality to less than 70 per 100,000 live births (UN, 2015). This shows the magnitude of ground to be covered to reduce the figure from 362 per 100000 live births in Kenya (Kenya National Bureau of Statistics et al., 2015). Efforts to improve maternal and neonatal health (MNH) in Kenya have concentrated on programs targeting the prenatal, perinatal and postnatal periods while little attention has been given to the preconception period, which is an essential element of the continuum of care. These programs as informed by the Kenya maternal and newborn health model are;-focused antenatal care, essential obstetric care, essential newborn care, targeted post-partum care, post abortion care and family planning (MOH, 2009). The Kenya maternal and neonatal health model, which was derived from the global strategy of safe mother hood, excludes the preconception care. Little is known as to why components of preconception care have not been included except for family planning.

The third decennial International Conference on Population and Development (ICPD) held in Cairo in 1994 emphasized the importance of reproductive freedom. It developed a definition of reproductive health that included “access to appropriate health care services that enable women to go safely through pregnancy and childbirth and provide couples with the best chance of having a healthy infant (Boulet, Parker, & Atrash, 2006). The 4^th^ ICPD conference identified zero maternal deaths, zero unmet need for family planning and zero gender-based violence and other harmful practices like female genital mutilation which are all preconception interventions as their priority agenda for the next decade (United Nations Population Fund, 2019). Preconception cares is key in improving maternal and neonatal health indicators (Dean et al., 2014). It also creates demand for other services like antenatal care. The PCC implementation is yet to be measured in the current setting.

## METHODS

### Study design and study area

A descriptive and analytical cross-sectional study was carried out in facilities in Kisumu County. Kisumu County is one of the new devolved counties of Kenya. Its borders follow those of the original Kisumu District, one of the former administrative districts of the former Nyanza Province in western Kenya. Its headquarters is Kisumu City. It covers an area of 805 square miles, and has population of 968,879 (KNBS, 2009). It has 7 sub-counties, namely, Kisumu West, Kisumu Central, Kisumu East, Seme, Muhoroni, Nyando and Nyakach. Kisumu County has a total of 129 public health facilities.

Cross-sectional studies are used to assess the burden of disease or health needs of a population and are particularly useful in informing the planning and allocation of health resources. Since no similar studies have been done on this area it provided an opportunity to generate hypothesis. Further the objectives of this study did not require repeated responses and could be achieved with information taken at one point in time.

The prevalence of preconception care was measured using the Facility Checklist (additional file) which had a list of preconception care interventions whose availability was used to draw a conclusion. The checklist required a response of yes or no as to whether the particular services were being provided or not. The mean of the proportions of interventions provided was presumed to be the rate of preconception care provision.

### Sample size and sampling

A sample size of 28 facilities was derived from a sampling frame of 129 health facilities using the corrected Fishers formula (Fisher et al., 1935). The study employed multistage sampling whereby stratified sampling was done first with the 7 sub-counties serving as strata ten in each subcounty KEPH levels formed the strata. Further, purposive sampling was used to select 2 high volume dispensaries, 1 health center, and 1 sub-county hospital from each sub-county

### Data collection

The study aimed at measuring the implementation of preconception care interventions in the afore mentioned County. Data was collected using a facility checklist (Additional file 1) that was based on a list of interventions identified as solutions addressing particular risk factors during preconception period that can influence pregnancy outcomes by a WHO consortium preconception care (Dean et al., 2013). The facility checklist had 15 service packages with several interventions under them that were observed for determination of PCC implementation in the facilities. All the interventions from all the service packs were adding up to 100, thus formed perfect denominator for percentage value determination of rate of provision. The fifteen service packs for PCC were as follows: Nutritional Management, Vaccination, Tobacco Use Prevention, Environmental Risk Exposure Reduction, Genetic Disorder Management, Planned Pregnancies, Sexually Transmitted Diseases Management, HIV Prevention and Management, Infertility and Sub-fertility management, Female Genital Cut Prevention and Management, Mental Health Disorders Management, Psychoactive Substance Use Prevention and Management, Intimate Partner and Sexual Violence Management, General Counselling and Diagnosing and Managing.

### Data management and statistical analysis

Proportions of the *yes* and *no* response, on whether each of the interventions for each service in the package as per the checklist was provided, were calculated and presented as percentages. Then, the means for all of the services in the package was determined. The significance of the difference in the means was determined by the one sample T-test at *P*-value of equal to or less than 0.05. This is because the sample size of 28 facilities met the criteria of this test statistic. Further the rate of use was derived from the proportion of women that each health provider indicated they had given care. The data was presented in form of tables and descriptions

## RESULTS

From the study, it was realized that the rate of PCC provision was quite low at 39%. The mean provision was observed to be lower in the primary level facilities (KEPH level 2 and 3) at 34% and higher in referral facilities (level 4 and above) at 45%. The service pack most provided at referral facilities was sexually transmitted infections management and general counselling service pack at 100 % (N=28). The poorly performing services were Environmental risk reduction at 27 % (N=28) and diagnosis and treatment of chronic diseases at 35% (N=28). The service with the highest implementation level was HIV prevention and management (84%) followed by sexually transmitted diseases (at 80%) and vaccination services (at 75%) in primary level facilities. The variation in prevalence per level of facility is further illustrated in **Figure 1**. The service with lowest level of implementation was environmental risk exposure reduction at 13% for level 2 and 3 followed by management of mental health disorders. Details on the rate of provision for the 15 care packages are described below and further profiled in **Table 1**.

**FIGURE 1.**
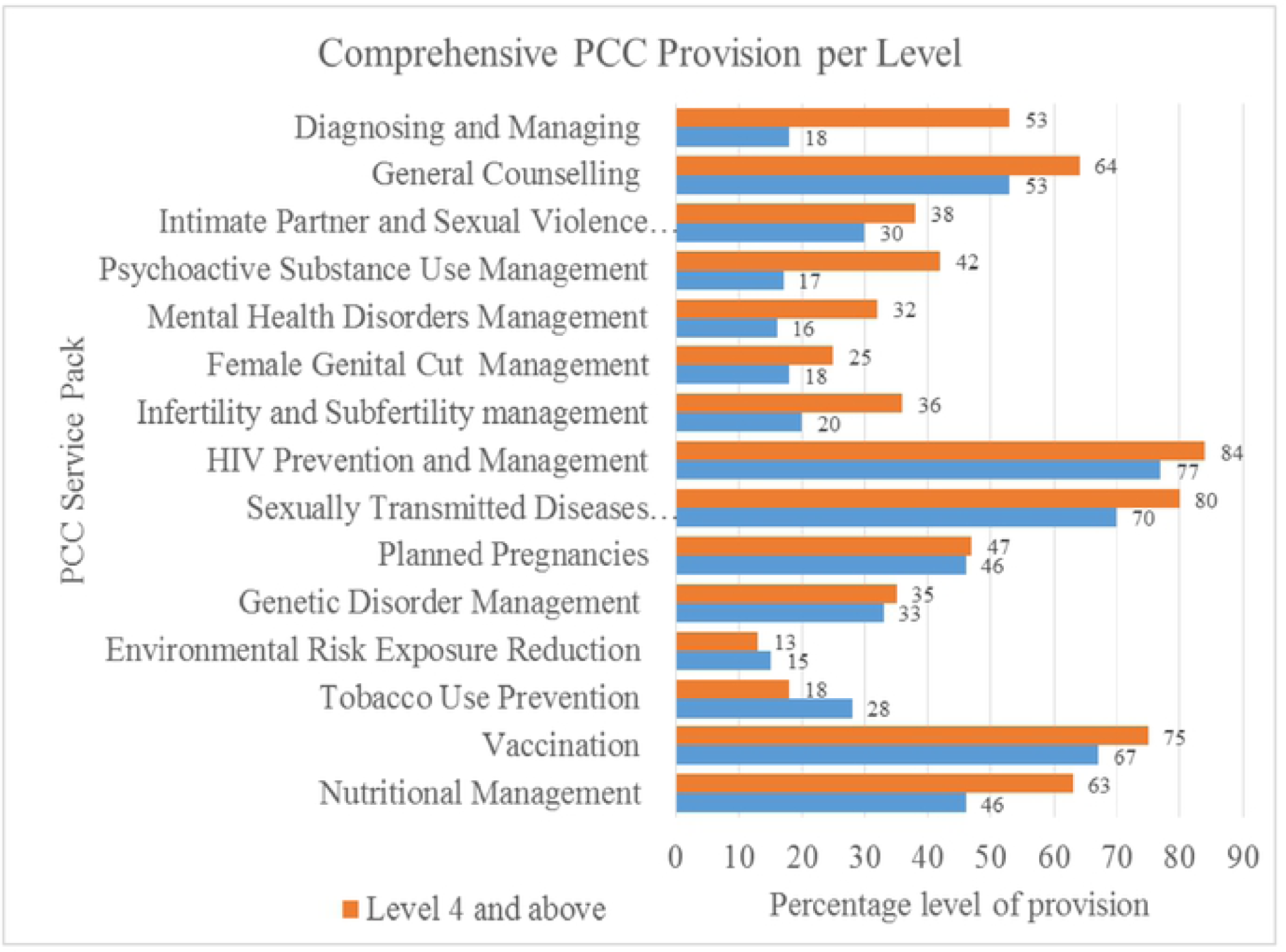
Rate if provision in Primary facilities (level 2 and 3) verses referral facilities(level 4 and above)

### Nutritional deficiencies and disorders management

Several interventions for the management of nutritional disorders and deficiencies were measured. They included screening for anemia, diagnosing and managing nutritional conditions, screening for diabetes, supplementing iron and folic acid, information, education and counseling, monitoring nutritional status, management of diabetes, including counseling people with diabetes mellitus, promoting exercise and iodization of salt. Level 4 and above facilities had the overall best performance (79%) in this area. The aspects that were implemented dismally were: promoting exercise and iodization of salt.

### Vaccination against Vaccine-preventable infections

The PCC care Package included vaccination against rubella, vaccination against tetanus and diphtheria and vaccination against Hepatitis B, which are the vaccines of high, impact on the outcome of the pregnancy. All facilities provided hepatitis vaccination while vaccination for tetanus and pertussis was more common in level 4 and above facilities. Rubella vaccine was only provided in some facilities across board.

### Prevention of tobacco use

Interventions to discourage tobacco use during preconception period were not implemented in Level 3 and below. In Level 4 and above, only screening of women and girls for tobacco use using “5 As” (ask, advise, assess, assist, arrange) was implemented at 7%. This accounted for approximately 1% of the intervention. This service pack contained other interventions including counseling on harmful effects of tobacco on pregnant women and unborn children, providing brief tobacco cessation advice, pharmacotherapy (including nicotine replacement therapy, if available), screening of all non-smokers and advising about harm of second-hand smoke and screening of women and girls for tobacco use.

### The environmental risks exposure reduction

The interventions to minimize exposure to environmental risks were; information on radiation exposure in occupational, environmental and medical settings, provide protection from unnecessary radiation exposure, information provision on avoiding unnecessary pesticide use, protecting from lead exposure, informing women of childbearing age about levels of methyl mercury in fish and promoting use of improved stoves and cleaner liquid/gaseous fuels. These environmental risk exposure interventions were not implemented at all in the various KEPH levels.

### The genetic disorders management

This package had several interventions that the prospective family has to be taken through as PCC. They included taking a thorough family history to identify risk factors for genetic conditions, family planning, genetic counseling, carrier screening and testing and appropriate treatment of genetic conditions. Only family planning intervention was provided at 100% at all levels.

### Prevention of early, unwanted and rapid successive pregnancies

Several interventions aimed at prevention of unwanted pregnancies were implemented at level 4 and above alone. These were: keeping girls in school initiatives (7%), engaging men to critically assess norms and practices regarding gender-based violence and engaging men to critically assess norms and practices regarding coerced sex (13% each). Other key aspects like Influencing cultural norms that support delayed marriage and consensual sex and empowering girls to resist coerced sex were not implemented at all.

### Management of sexually transmitted infections

The management of sexually transmitted infection (STI) was generally well implemented, several at 100% across all KEPH levels. The interventions were: diagnosing and treating STIs e.g., syphilis, gonorrhea and genital ulcer diseases, promoting condom use for dual protection against STIs and unwanted pregnancies, ensuring steady access to condoms and screening for STIs. Equally, promoting safe sex practices through individual, group and community-level interventions and STIs treatment and other relevant health services performed highly at 92% and 100% for lower and higher KEPH levels respectively. Only one service, providing age-appropriate comprehensive sexuality education and services, performed dismally.

### HIV-AIDS prevention and management

Seven of the eight services for the management of HIV were at optimum performance of 100%. These 7 were: family planning, provider-initiated HIV counseling and testing, including male partner testing, providing antiretroviral treatment (ART) for prevention of mother to child transmission of HIV, providing ART for pre-exposure prophylaxis, providing ART for post-exposure prophylaxis, providing male circumcision and determining eligibility for lifelong antiretroviral therapy. Only promoting safe sex practices and dual method for birth control and STI control was at 92% for Level 3 and below and 75% for level 4 and above.

### Management of fertility and sub-fertility

The management of fertility and sub-fertility package had 7 interventions. Three of the seven, namely, defusing stigmatization of infertility and assumption of fate, counseling on infertility and counseling for those diagnosed with unpreventable causes of infertility/sub-fertility were not observed at all the levels. Screening and diagnosis of couples following 6–12 months of attempting pregnancy and management of underlying causes of infertility/sub-fertility, including past STIs were implemented at both levels at 9% for both for Level 3 and below and 25% and 32% for level 4 and above, respectively.

### Prevention and management of female genital mutilation (FGM)

There were 5 interventions under FGM PCC service pack. These were: hosting public forums on discussing and discouraging the FGM, screening women and girls for FGM to detect complications, informing women and couples about complications of FGM and access to treatment, providing corrective surgery for women with complications of FGM and removing cysts and treating other complications of FGM. Only one out of the five, providing corrective surgery for women with complications of FGM, was implemented at 7% in the Level 5 Hospital.

### Mental health disorders management

Mental health intervention package had the following aspects; - providing general mental health education before pregnancy, providing psychosocial counseling before pregnancy, counseling and treating depression in women of childbearing age, strengthening community networks for women, promoting women’s empowerment, improving access to education and or information for women of childbearing age and reducing economic insecurity of women of childbearing age. Only counseling and treating depression in women of childbearing age (13%) and strengthening community networks for women (57%) were implemented in the level four and five hospitals.

### Psychoactive substance use prevention and management

Five out of the 9 interventions of the psychoactive substance use service pack were implemented in level 4 and 5 hospitals. These were: providing brief interventions and treatment when needed (38%), establishing prevention programs to reduce substance use in adolescents (7%), treating substance use disorders, including pharmacological (7%), providing psychological interventions for substance use (7%) and providing family planning assistance for families with substance use disorders (44%) and it was also implemented in the lower-level facilities at rate of 42%. The rest of the aspects of mental health: screening for substance use, changing individual and social norms regarding drinking, screening and counseling of people who are problem drinkers and treating people who have alcohol use disorders were not observed to be implemented in all the facilities.

### Intimate partner and sexual violence management

The intimate partner and sexual violence service pack had 7 interventions. Four of the 7 aspects were implemented in higher KEPH levels and 2 were implemented across all the levels. Those that were implemented were: health promotion to prevent intimate partner violence, and providing age-appropriate sexuality education that addresses human rights at 7% only in level 4 and 5 facilities, while recognizing signs of violence against women 9% and 25% and providing post-rape care, referral and psychosocial support to victims of violence 100% and 44% in level 3 and below and 4 and above respectively. The others were not implemented, namely: providing age-appropriate sexuality education that addresses gender equality, providing age-appropriate sexuality education that addresses sexual relations and linking economic empowerment, gender equality and community mobilization activities.

### General Counselling

Counselling on postnatal visits and about HIV/AIDS were the ones mostly done at 100% followed by counselling on recommended 4 ANC visits and birth attendance at 67%. Counselling about breastfeeding, and use of nets was rarely done at 17% while new-born care counselling was seldom done at 0% for the facilities visited.

### Diagnosis and management of specific diseases

This service pack was being poorly implemented at the facilities in lower KEPH levels (2%). There was no provision of diagnosis and management of rubella at all levels. The other aspects of diagnosis and management were implemented in level 4 and above as represented.

## DISCUSSION

Preconception care remains a key component in the continuum of maternal and neonatal health. This study realized that there was no specific PCC program. The overall level of provision of the various interventions that make up the preconception package was 39%. This was considerably high compared to other studies done in Africa. A study done in Ethiopia estimated it at 15% (Kassa et al., 2019). The sample size of this study was larger (n=634) which could have increased the precision of the findings. Secondly, to measure implementation the study administered questionnaires to health providers while the current study sampled facilities and ad a checklist filled.

These services were being provided individually and not as a package as recommended by the WHO. This is the lack of standardization of care, an issue that was also highlighted in a previous study (Lassi et al., 2014). Further a fragmented health care service delivery system, by way of lack of a comprehensive preconception care program, is a barrier to uptake of preconception care (M’Hamdi et al., 2017). Nevertheless, this ability to offer these services in isolation is an opportunity to create a sustainable comprehensive program. This is so because, other scholars have noted that causes of unhealthy birth growth and development are interwoven and that addressing them one at a time can solve only a small fraction of the problem (Dubei, 2014). Moose et al, 2008 recommended routine care to maximize on the gains of preconception care for better maternal outcomes (Moose & Cefalo, 2008). In Africa, South Africa have included it as a routine in the continuum of maternal care (Republic of South Africa, 2015).

Level of provision was observed to be lower in the primary level facilities (KEPH level 2 and 3) at 34% and higher in referral facilities (level 4 and above) at 45%. This can be due to the fact that referral facilities have most cadres of health workers including doctors and previous studies have shown that doctors demonstrated a higher likelihood to provide care than the other cadres (Genius & Genius, 2016). It could also be, because the referral facilities have better infrastructure and more specialised equipment that can make it possible to provide more services than the lower facilities. But this may not be favourable since a previous study revealed that women who received preconception care in either a healthcare centre or the community developed the appropriate behavioural changes as a result of the preconception care provided (Dean et al., 2014).

The service with the highest implementation level was HIV prevention and Management 84% followed by sexually transmitted diseases at 80% and vaccination services at 75%. These are services targeting all persons including women in childbearing to achieve other objectives rather than preconception care. Thus, it may imply that the rate of preconception maybe even lower than currently estimated. The service with lowest level of implementation was environmental risk exposure reduction at 13% for level 2 and 3 followed by management of mental health disorders.

## Conclusion and recommendation

This study has been able to establish the prevalence of PCC to be relatively low. More so in service that are not provided with any other objectives like environmental risk exposure and treatment of mental disorders. Thus, we recommend that any entity intending to increase provision of PCC should first focus on advocating for such services to improve their implementation while promoting integration. Further, we recommend studies to further profile the determinants of preconception care provisions in various levels of health care delivery.

## Data Availability

the data has been deposited in fig share and the DOI is 10.6084/m9.figshare.19596553

https://figshare.com/s/3cd8116bb22e76bb98cc

## Authors’ contributions

ENM designed, carried out the survey study and participated in the drafting of the manuscript. CO designed the study and participated in the drafting of the manuscript. ENM, MSS and CO performed the statistical analysis. All authors read and approved the final manuscript.

## Acknowledgements

We are indebted to the health facilities managers within the study settings in Kisumu County and the enumerators for data collection. These data are published with the approval from the MMUST Ethics and Research Committee and JOOTRH Ethics and research committee. This work is a continuation of ENM’s P h d in Public Health studies at Masinde Muliro University of Science and Technology, Kenya and received some funding support from the institution.

